# Based on breast CEUS parameters combined with serum CA153, a nomogram model was constructed to predict the molecular classification of breastcancer

**DOI:** 10.1101/2024.10.17.24315659

**Authors:** Yanlei Ji, Zhen Han, Yong Zhang, Wenwen Sun

## Abstract

**Objective:** To analyze the role of contrast-enhanced ultrasound (CEUS) parameters combined with serum tumor marker CA153 in the prediction of Breast Cancer (BC) molecular typing.

**Methods:** From January 2020 to January 2023, 120 BC patients diagnosed in our hospital were studied. According to the pathological results, the patients were divided into Luminal and non-Luminal BC groups. Both groups underwent contrast-enhanced ultrasoun. The time-intensity curve (TIC) is obtained, and the relevant characteristic parameters are obtained, including peak intensity (PI), peak time (TTP), area under the curve (AUC), and mean transit time (MTT). Serum tumor marker CA153 was detected in both groups. Combined with CEUS characteristic parameters and serum CA153 of two groups of BC patients, a multiple Logistic regression model was constructed, and a nomogram prediction model was constructed based on the model. Calibration curve and receiver operating characteristic (ROC) curve were used to analyze the value of this model in the prediction of BC molecular classification.

**Results:** There were no significant differences between Luminal BC patients and non-Luminal BC patients in clinical parameters and qualitative parameters of contrast-enhanced ultrasound, while there were statistical differences between quantitative parameters PI, AUC and serum tumor marker CA153. The AUC of the combined diagnosis of three parameters (PI, AUC and CA153) was significantly higher than that of the single index diagnosis group. The ROC curve AUC of BC molecular typing was predicted to be 0.94 based on the three-parameter nomogram, and the fitting of the actual curve and the ideal curve in the calibration curve was close.

**Conclusions:** The nomogram model based on breast contrast-enhanced ultrasound (CEUS) parameters combined with serum CA153 can effectively predict the molecular classification of BC.

## Introduction

Breast Cancer (BC) is one of the most common malignant tumors in women. In recent years, its incidence has been increasing year by year and showing a trend of youth, and seriously threatening the life health and quality of life of patients^[1]^. BC is a hormone-dependent tumor, and its carcinogenic risk is closely related to estrogen imbalance^[2–4]^. Estrogen is mediated by estrogen receptor (ER) and progesterone receptor (PR) and its expression level is different in dif ferent BC patients, and its clinical manifestations, treatment and prognosis are also different^[5, 6]^. Based on the expression status of ER and PR, BC is divided into Luminal type (positive for either ER or PR, including Luminal A and Luminal B) and non-Luminal type (negative for both ER and PR, including HER-2 and TN)^[7]^. Therefore, the development of accurate diagnostic methods for B C patients with different molecular subtypes is very important for the treatment and prognosis of patients^[8]^. In recent years, with the development of molecular biology technology, more tumor molecular markers related to BC have been discovered. Evaluating the expression levels of these markers in the serum of patients can provide valuable information for the occurrence, invasion and metastasis of BC^[9]^. Studies have suggested that carbohydrate antigen 153 (CA1563), carcinoembryonic antigen (CEA) and carbohydrate antigen 125 (CA125) have certain clinical value for BC screening, postoperative follow-up, recurrence and metastasis, especially CA153, which is the most valuable single diagnostic index with the best sensitivity, accuracy, positive predictive value and specificity^[10]^. However, CA153 alone cannot accurately reflect the overall situation of the les ion. contrast-enhanced ultrasound (CEUS), as a non-invasive imaging technique, can observe the microcirculation in BC lesions. At the same time, it can fully reflect the information of the internal and peripheral tissue structure of the lesion^[11]^. Based on this, the present study constructed a nomogram diagram with CEUS parameters combined with serum tumor marker index CA153 to predict different BC molecular typing, with a view to providing clinical reference for precise diagnosis and personalized treatment of BC.

## Materials and methods

### 1.1 Patients

01/09/2024 searched the HIS information of our hospital, and selected 120 breast cancer patients diagnosed by examination from January 2020 ∼ January 2023 as the research subjects. All patients underwent ultrasonography and clinical immunohistochemical pathology. Inclusion criteria: Invasive BC was confirmed by needle biopsy and immunohistochemical evaluation was performed; No chronic diabetes mellitus; Patients volunteered to participate in the study; Complete medical records. Exclusion criteria: serious cardiovascular and cerebrovascular diseases and digestive tract diseases; Hypertension and autoimmune disease; Pregnancy or breastfeeding; Allergic to ultrasound contrast agent. This study was approved by the Ethics Committee of our hospital, and all patients have signed informed consent.

### 1.2 Contrast-enhanced ultrasound

GE Logiq E9 color Doppler ultrasonic diagnostic instrument, ML6-15 probe, frequency 9∼15 MHz; Equipped with contrast-enhanced ultrasound imaging software. First, routine ultrasound was performed to observe the conditions of both mammary glands and determine the location of breast lesions to obtain better two-dimensional images. The optimal section was selected to cut into the angiography state, and 2.4 ml Sonovi (Bracco, Italy) was injected rapidly through the cubitus vein, and then rinsed with 5.0 ml 0.9% normal saline. The dynamic perfusion process of breast lesions was observed and recorded. The region of interest (diameter 4 mm, area 8 mm^2^) was selected when the breast lesions reached the peak of enhancement, and 5 points were selected from the inside and the edge of the lesions as the region of interest. The measured values of the region of interest at a total of 10 points were recorded and their mean values were taken. Start the QLAB analysis software to automatically analyze and obtain the time-intensity curve (TIC), and perform curve fitting to obtain the blood flow parameters of the region of interest: peak intensity (PI), peak time (TTP), area under the curve (AUC) and mean transit time (MTT). The above images were analyzed by two physicians. If there were any differences in the diagnosis results, the third physician would participate in the discussion and get the final result after reaching an agreement.

### 1.3 Immunohistochemical examination

The expression levels of ER, PR, HER-2 and Ki-67 were detected in all BC samples after paraffin embedding and SP staining. ER, PR Rubric^[12]^: ER and PR positive tumor nuclei ≥1% positive, < 1% negative. HER-2 Rubric^[13]^: Negative 1+ is HER-2 negative and 3+ is HER-2 positive; those with 2+ need to be tested by fluorescence in situ hybridization, in which those with gene amplification are HER-2 positive and those without amplification are HER-2 negative. Ki-67 proliferation index^[12]^: Select the area with the highest staining density of tumor cells, and count the staining of 1000 cells. The number of positive cells/total cell count is the Ki-67 proliferation index.

### 1.4 CA153 detection and judgment criteria

2 mL of fasting venous blood was collected from both groups of patients in the early morning, and then centrifuged at 3000 r/min for 10 min after self-coagulation to separate the serum, which was stored in the refrigerator at -20℃ for examination. Serum CA153 concentration was detected by using Abbott Architecti 2000 automatic chemiluminescence immunoassay analyzer and special reagents. All operations were performed in strict accordance with the instrument and reagent instructions.The critical value of CA153 was 31.3 U/mL, and the assay was considered positive if the value exceeded the critical value.

### 1.5 Data analysis

All analyses were performed using R software version 4.0.2. Logistic regression and ROC analysis using R software. Measurement information data were expressed as Mean±SD and one-way ANOVA was used for comparison between groups. Count data data were expressed as frequencies and χ^2^ test was used. Receiver operating characteristic curves (ROC) were used to analyze the diagnostic performance of the model for different BC subtypes. *P*<0.05 was statistically significant.

## Results

### 2.1 Clinical characteristics

The 120 study patients ranged in age from 24 to 80 years. There were 72 patients with Luminal BC and 48 patients with non-Luminal BC. The general data of the study patients included age, maximum mass diameter, menopause, pathological type, and pathological grade, and the results were shown in Table 1. There was no significant difference in the general data of 120 patients with different BC types.

**Table 1.**
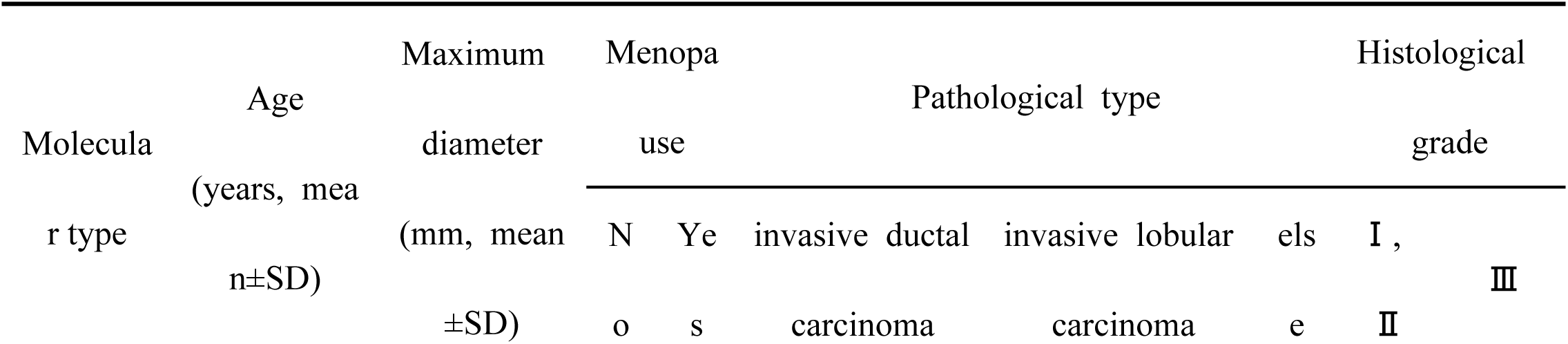
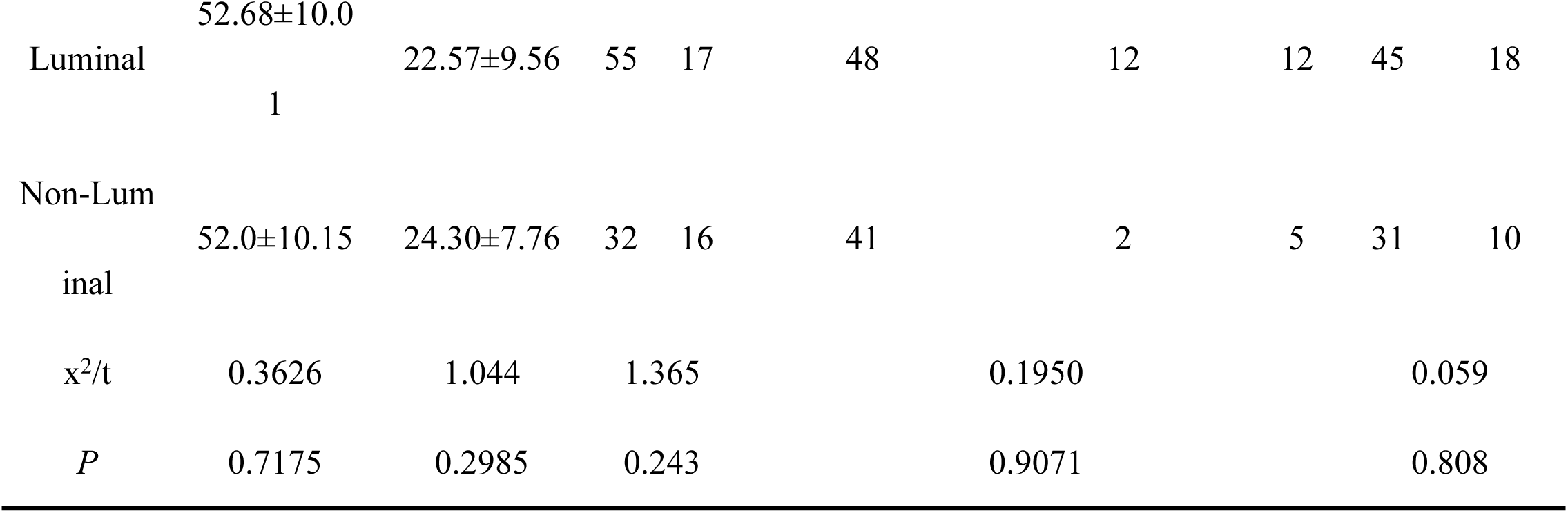
Clinical characteristics.

### 2.2 CEUS manifestations

The results of comparing the qualitative parameters of ultrasonography in patients with different BC subtypes are shown in Figure 1 and Table 2. In patients with the Luminal type, ultrasonography is dominated by centripetal hyperenhancement, with a predominantly rapid ascending and slow descending enhancement pattern, and unclear and enlarged enhancement with radiolucent convergence of the borders; Centripetal enhancement accounted for 14/39 patients with the non-Luminal type, with a predominantly rapid ascending and slow descending pattern of enhancement, unclear and enlarged borders after enhancement, and radiolucent convergence of the borders.

**Figure 1.**
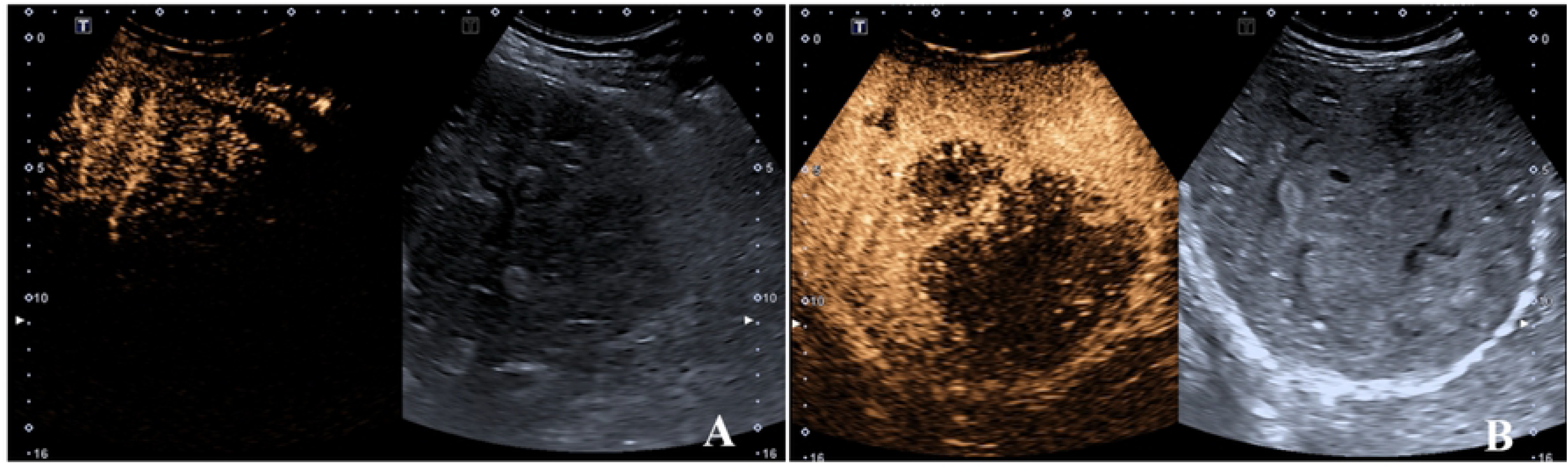
Contrast-enhanced ultrasound of different molecular types of BC. (A: Ultrasonography of a patient with Luminal B BC showing radial convergence around the periphery of the mass on ultrasonography; B: Ultrasonography of a patient with TN BC showing a well-defined and well-formed lesion on ultrasonography)

**Table 2.**
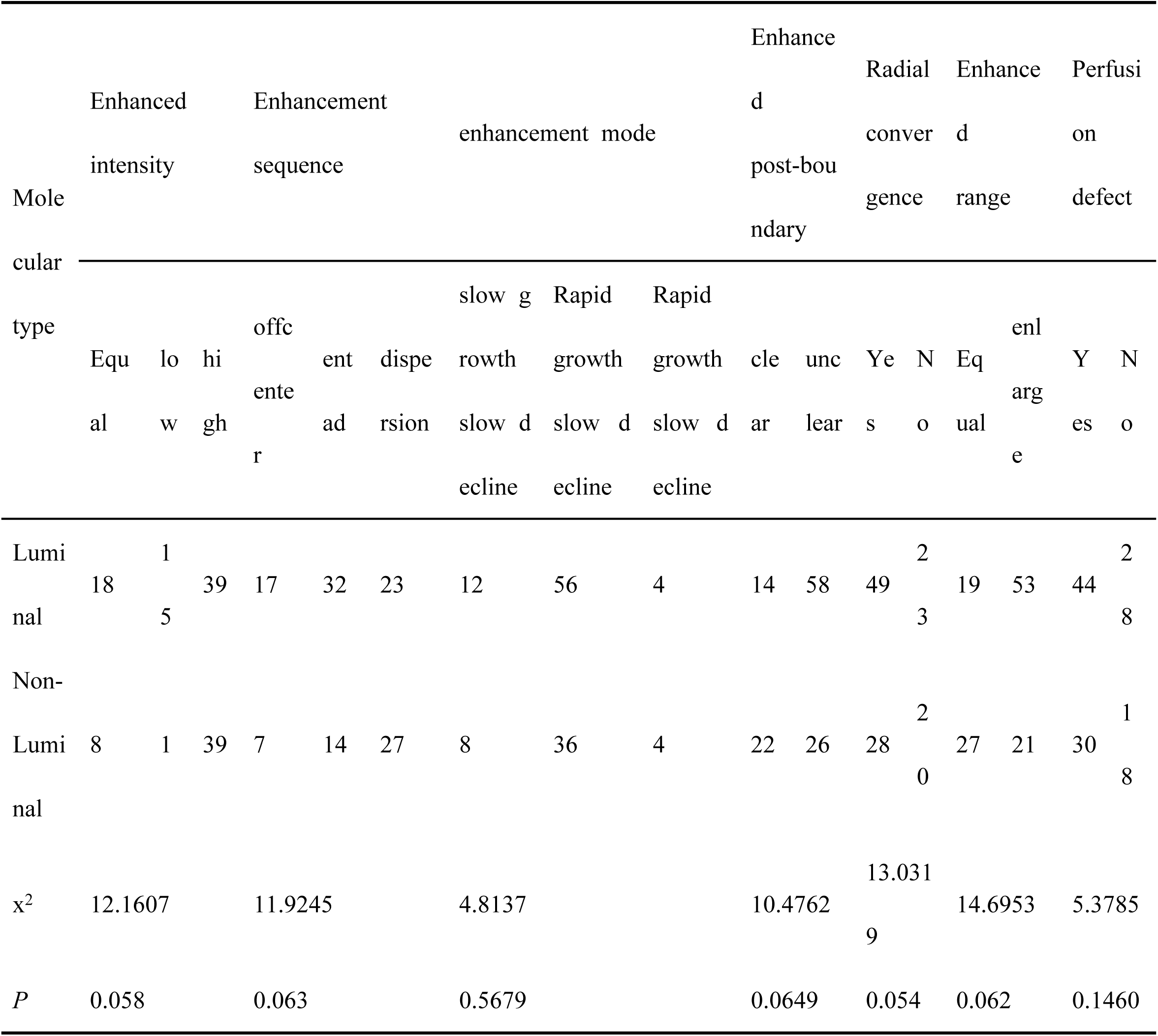
Contrast-enhanced ultrasound findings of patients with different BC types.

### 2.3 Comparison of CEUS characteristic parameter and CA153

The CEUS quantitative parameters and serum CA153 indexes of patients with different BC types were compared, The results are shown in Table 3. There were significant differences in PI, AUC and serum CA153 between Lumina l and non-Luminal BC patients.

**Table 3.**
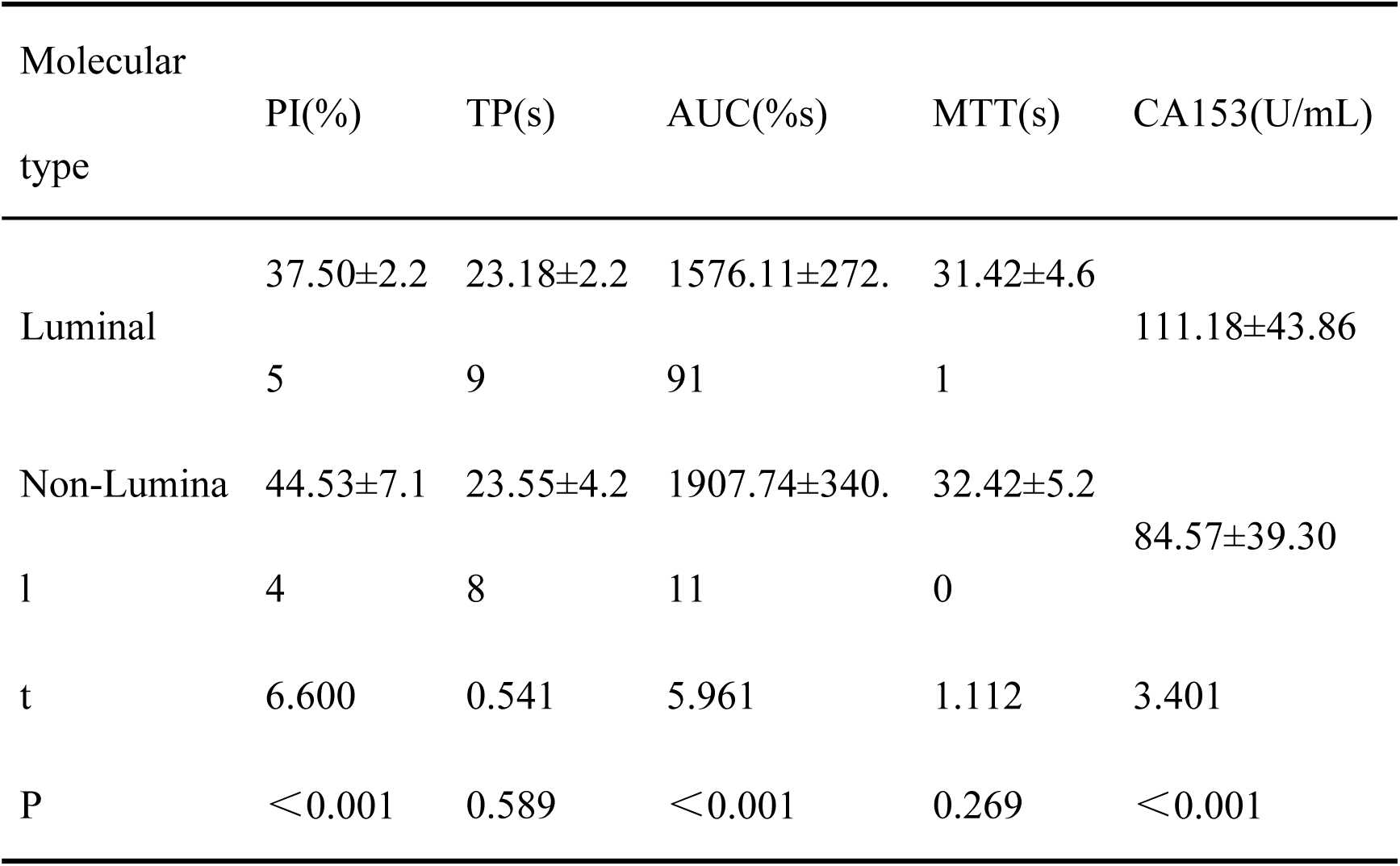
Comparison of CEUS characteristic parameter and CA153.

### 2.4 Binary logistic regression analysis screened the independent predictors of BC molecular typing

To further evaluate the predictive performance of serum tumor marker CA153 and contrast-enhanced ultrasound parameters in different types of BC. Univariate regression analysis was performed to compare the clinical data, CEUS quantitative parameters and serum tumor marker CA153 between Luminal and non-Luminal subtypes. Subsequently, univariate factors with P values < 0.05 were included in the multivariate regression analysis, including CEUS parameters PI, AUC and serum tumor marker CA153. Multivariate logistic regression analysis showed that PI, AUC and CA153 were independent predictors of BC molecular typing (P < 0.05).(Table 4).

**Table 4.** Binary logistic regression analysis screened the independent predictors of BC molecular typing.

| Variate | Univariate regression analysis |  |  | Multivariate regression analysis |  |  |
| --- | --- | --- | --- | --- | --- | --- |
|  | OR | 95%CI | P | OR | 95%CI | P |
| Age | 0.99 | 0.96,1.03 | 0.715 | - | - | - |
| Menopause | 1.47 | 0.65,3.33 | 0.355 | - | - | - |
| Maximum diameter | 1.02 | 0.98,1.07 | 0.297 | - | - | - |
| Histological grade | 0.79 | 0.33,1.90 | 0.597 | - | - | - |
| Enhanced intensity | 2.25 | 0.88,5.78 | 0.092 | - | - | - |
| Enhancement sequence | 2.73 | 0.97,7.71 | 0.058 | - | - | - |
| enhancement mode | 1.50 | 0.29,7.81 | 0.630 | - | - | - |
| Enhanced post-boundary | 0.29 | 0.13,0.64 | 0.103 | - | - | - |
| Radial convergence | 1.52 | 0.71,3.25 | 0.278 | - | - | - |
| Enhanced range | 0.58 | 0.13,0.60 | 0.215 | - | - | - |
| Perfusion defect | 0.99 | 0.96,1.03 | 0.087 | - | - | - |
| PI | 1.36 | 1.20,1.53 | < 0.001 | 1.31 | 1.09,1.58 | 0.004 |
| TP | 1.04 | 0.92,1.16 | 0.542 | - | - | - |
| AUC | 1.01 | 1.01,1.01 | < 0.001 | 1.01 | 1.01,1.01 | 0.002 |
| MTT | 1.04 | 0.97,1.13 | 0.269 | - | - | - |
| CA153 | 0.98 | 0.98,0.99 | 0.002 | 0.98 | 0.96,0.99 | 0.001 |

### 2.5 Construction of nomogram prediction model and comparative analysis of prediction efficiency

Based on the independent predictors (PI, AUC, CA153) of the three molecular subtypes of BC screened by binary logistic regression analysis, ROC curve was established to evaluate the accuracy of the combination of the three parameters in predicting non-Luminal BC (Figure 1). The results showed that the area under the curve (AUC) of the three-parameter model was 0.8307 (95%CI: 0.7437-0.9177), the sensitivity and specificity of the prediction were 75% and 94.44%, respectively, and the cut-off value of the prediction model was 0.3437.This indicates that the three-parameter combined prediction model of PI, AUC and serum CA153 has good predictive performance for the diagnosis of non-Luminal events (Table 5). Finally, we constructed a nomogram to predict the probability of non-Laminal BC based on the PI, AUC, and CA153 mentioned above (Figure 2). Each independent variable value corresponds to a score in the top row, and the scores of the individual independent variables are summed to obtain the total score, from which the probability is subsequently calculated. Finally, we used ROC curve and calibration curve to evaluate the predictive performance of the nomogram (Figure 3,4). The AUC of ROC curve was 0.94 (95%CI: 0.90-0.98). In the calibration curve, the actual curve was very close to the ideal curve. The nomogram has good predictive value for predicting the risk of non-Luminal BC.

**Figure 2.**
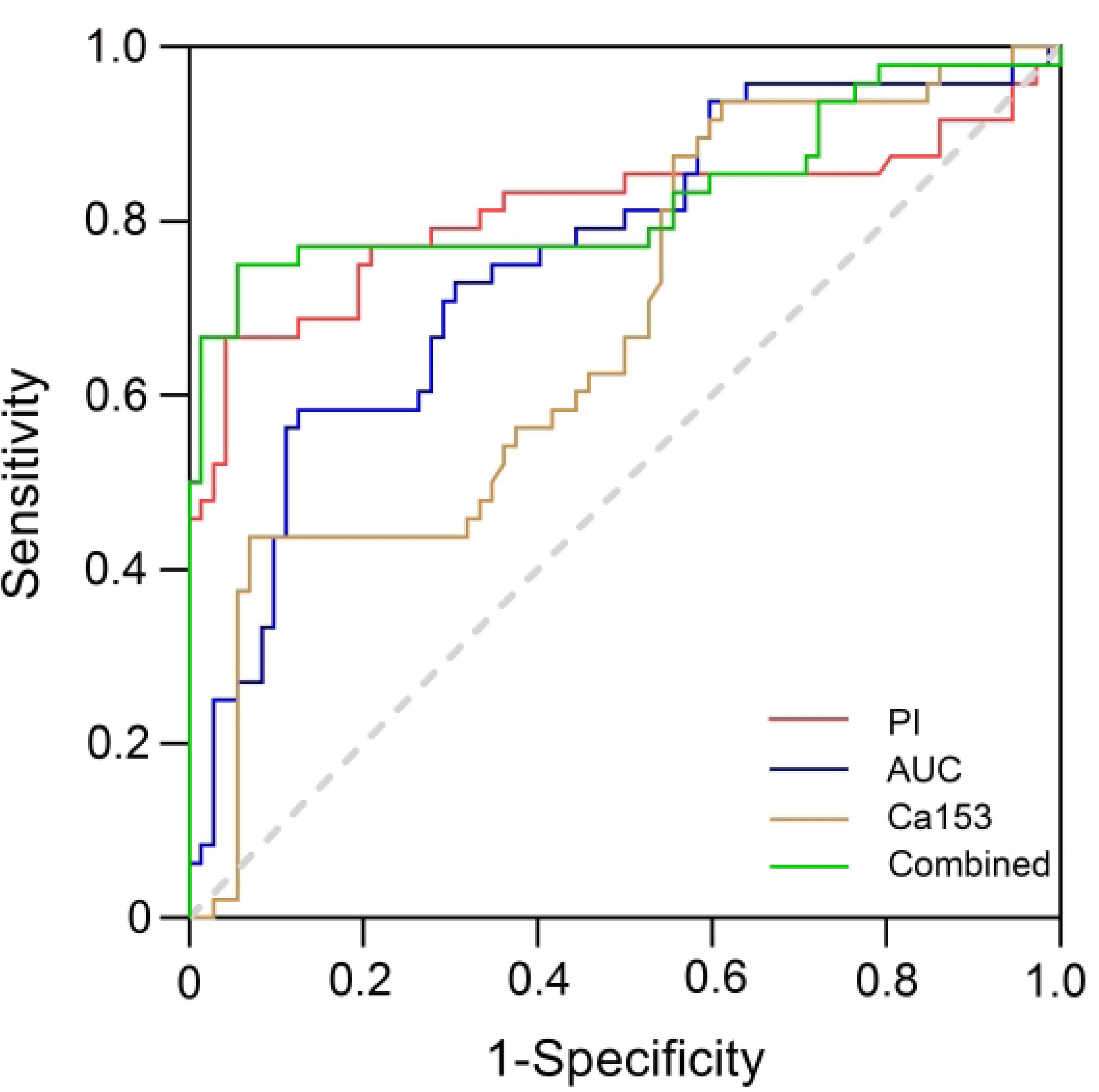
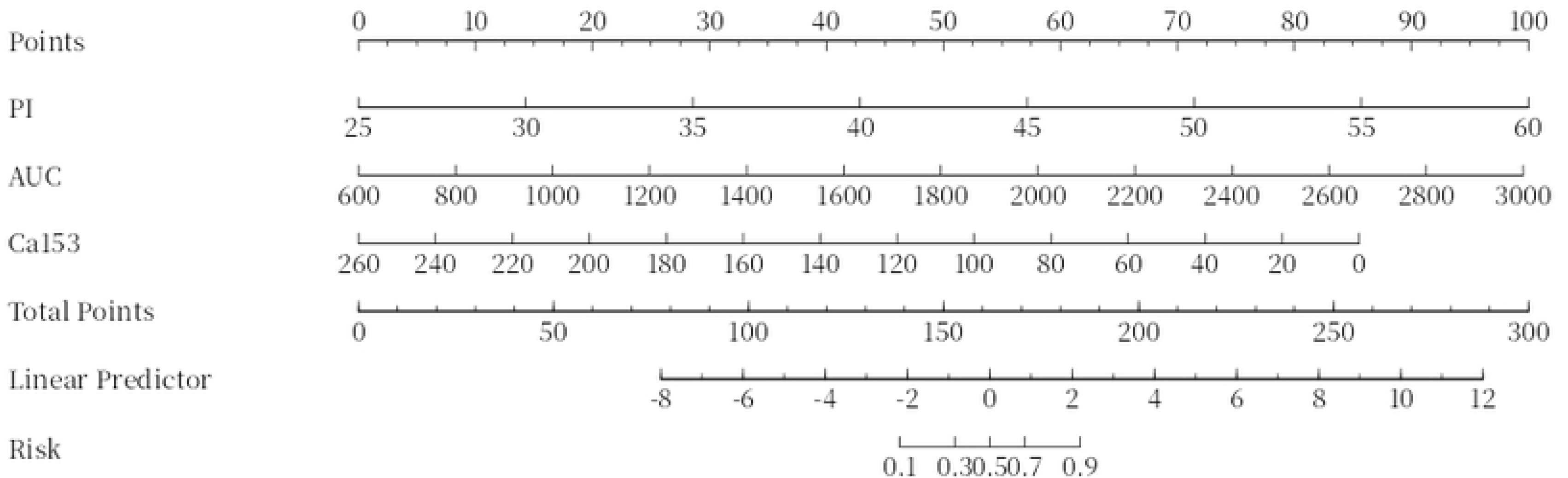
**ROC curve was predicted by three-parameter combination**

**Figure 3.**
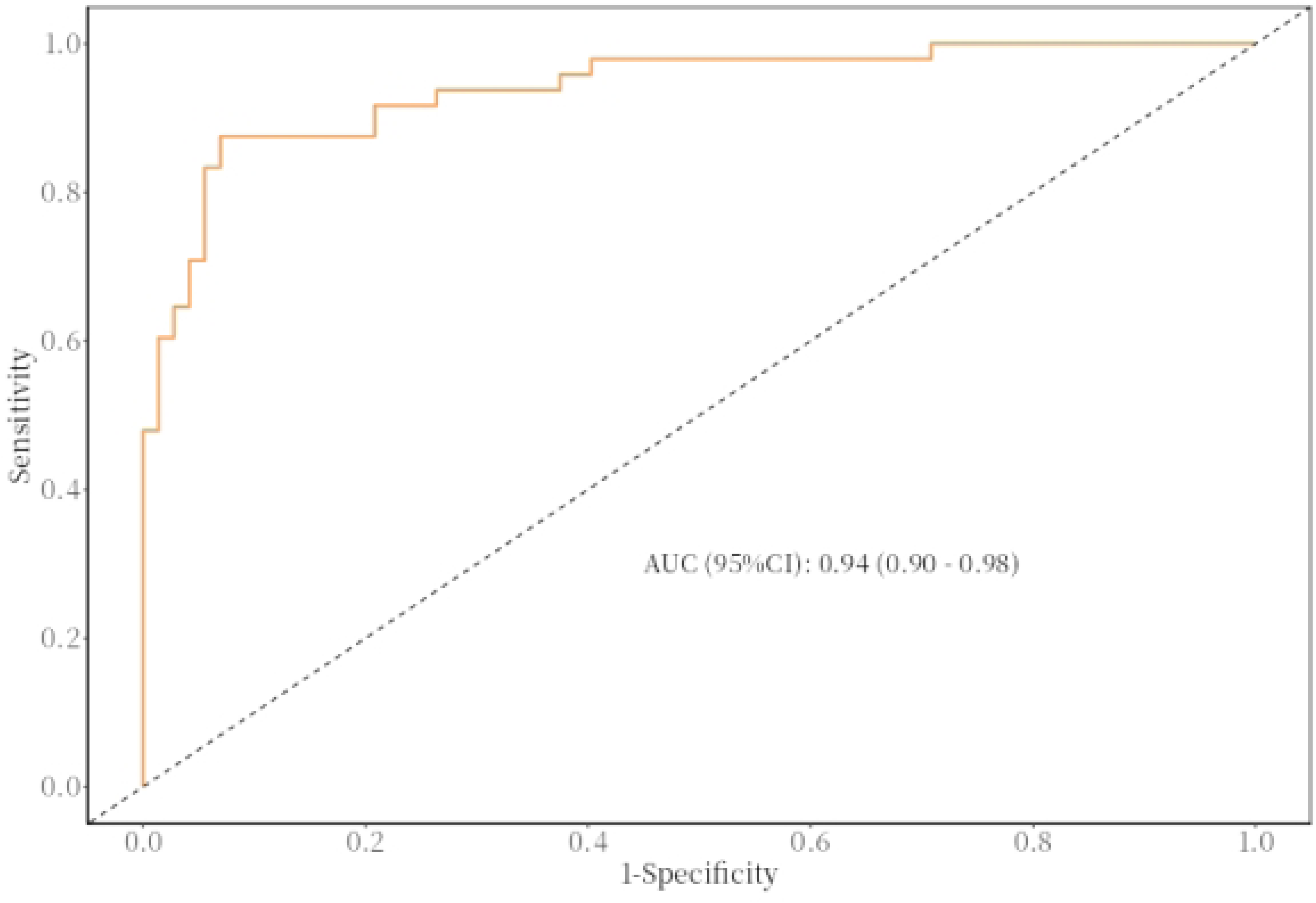
**ROC curve of the nomogram model**

**Figure 4.**
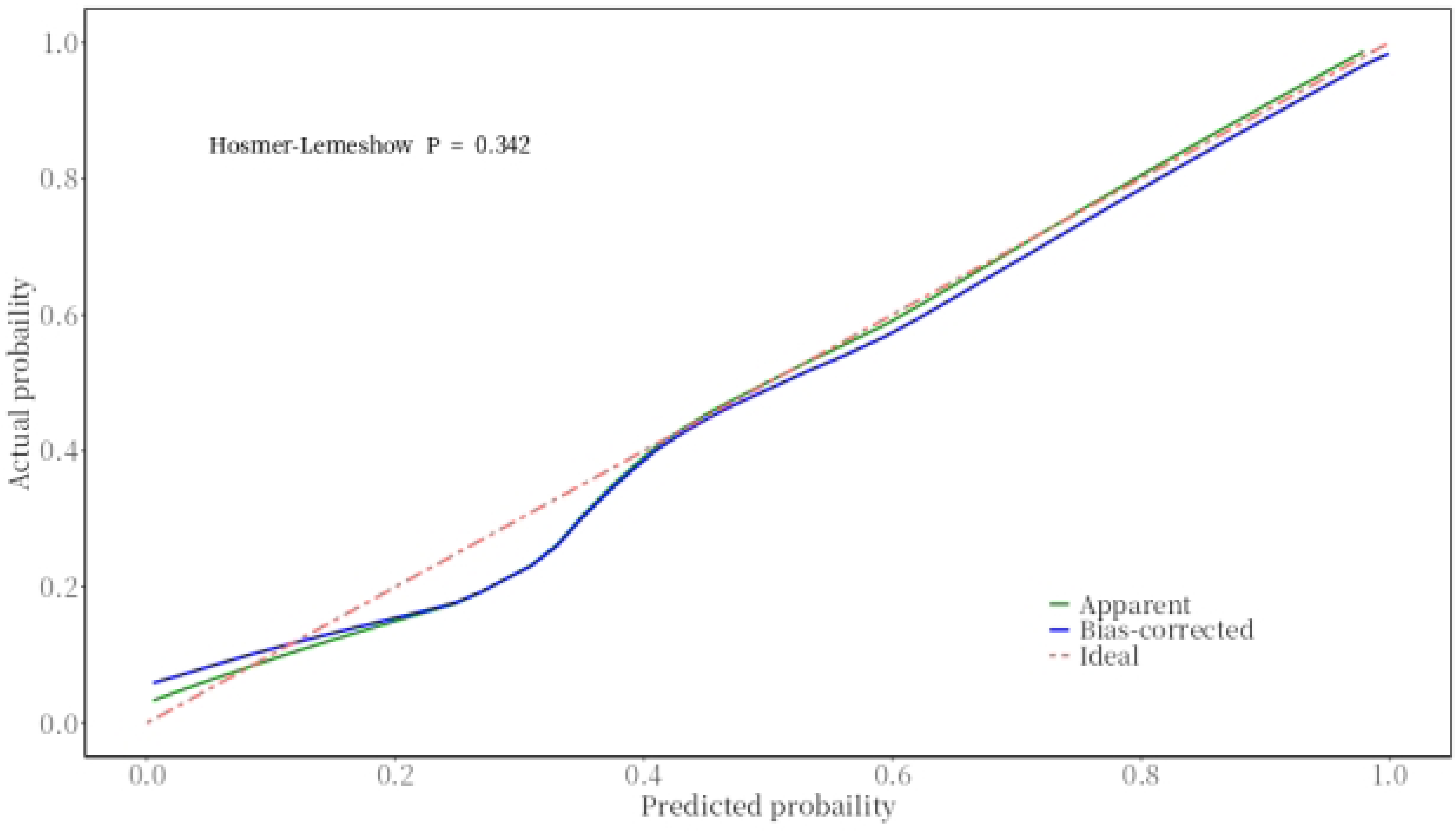
**Calibration curve of nomogram model**

**Table 5.**
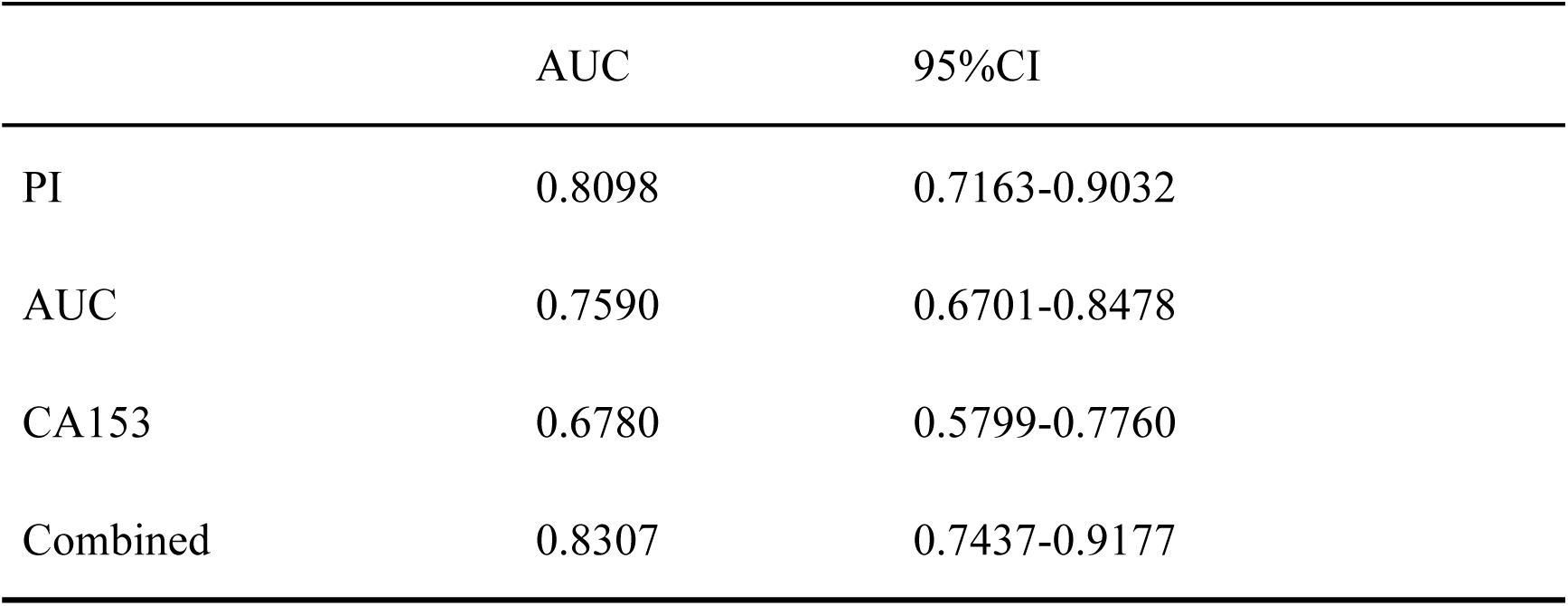
Comparison of AUC values.

|  | AUC | 95%CI |
| --- | --- | --- |
| PI | 0.8098 | 0.7163-0.9032 |
| AUC | 0.7590 | 0.6701-0.8478 |
| CA153 | 0.6780 | 0.5799-0.7760 |
| Combined | 0.8307 | 0.7437-0.9177 |

## Discussion

BC is a highly vascular heterogeneous and malignant stromal tumor, and i ts incidence is increasing year by year, which has become the first disease seri ously endangering women’s health^[14]^. The etiology of the disease is still unclear, and genetic factors and estrogen endocrine abnormalities are the main causes of the disease^[15]^. In 2000, Perou et al^[16]^ first proposed that BC has different molecular subtypes, which can be divided into Luminal type and non-Luminal type (including HER-2 type and TN type). Studies have found that different molecular subtypes of BC have different biological behaviors, chemotherapy sensi tivity and prognosis. Among them, Luminal BC is the most common molecular subtype, accounting for about 50%-70% of BC including Luminal A and Lumi nal B subtypes. Non-Luminal BC includess two different subtypes, HER-2 and TN subtypes, and this molecular subtype of BC is highly heterogeneous and characterized by high invasiveness. Compared with Luminal BC, it is more difficult to diagnose and treat. Therefore, early detection, early diagnosis and early treatment of different molecular subtypes of BC are of great significance for improving the survival rate and quality of life of patients.

The occurrence, development, invasion and metastasis of BC are dependent on neovascular vessels. The marginal zone of tumor is an area with vigorous growth of tumor cells. The blood vessels are dense and densely distributed, anastomosing with each other to form loops, and some blood vessels are dilated in sinus shape^[17]^. CEUS, as a non-invasive imaging method for preoperative evaluation of tumor angiogenesis, can accurately display blood perfusion compared with conventional ultrasound. The application of contrast-enhanced ultrasound c an not only clearly observe the course and distribution of microvessels in the tumor, but also provide quantitative analysis of the tumor, providing imaging reference for the preoperative diagnosis and treatment of BC^[18]^. In this study, we compared the characteristics of contrast-enhanced ultrasound between the two groups of patients. We found that the contrast-enhanced ultrasound of Luminal B C showed high enhancement, unclear boundary, radial convergence around the periphery, and expanded range after enhancement, and the enhancement pattern was mostly rapid increase and slow decrease. This result is the same as that o f Wang Xiaoyan et al^[19]^, and can be used as a sensitive sign of Luminal BC. Non-Luminal BC showed high enhancement on contrast-enhanced ultrasound, an d the boundary was basically unclear after enhancement, and the boundary did not significantly expand after enhancement, similar to the imaging characteristics of benign tumors, which was rare in other molecular subtypes, highlighting t he heterogeneity of this subtype. Whether the quantitative parameters of contrast-enhanced ultrasound are correlated with the molecular subtypes of BC, many studies have conflicting conclusions^[20]^. Lin Yun^[21]^ believes that there is a cert ain correlation between the quantitative parameters of contrast-enhanced ultrasound in different molecular types of BC, and Jia Wanru^[22]^ believes that there is no statistically significant difference in the quantitative parameters of contrast-enhanced ultrasound between different molecular types of BC. In this study, we found that there were significant differences in PI and AUC of contrast-enhanced ultrasound in patients with different BC subtypes, which may have guiding significance for predicting different molecular types of BC.

At present, with the rapid development of genomics and molecular biology, different molecular subtypes of BC can better reflect the biological behavior of tumors. The detection of serum tumor markers can not only assist in diagnosis, but also effectively predict the efficacy, outcome and acquired drug resistance. CA153 is an antigen closely related to human breast malignant tumors. It was first found in BC epithelial cells as a variant of the epithelial surface glycoprotein of breast cells. When a cell undergoes cancerous transformation, the activa tion of glycosyltransferases causes changes in cell surface sugars and enters the blood with the tumor cells^[23]^. It has obvious tumor specificity for BC and is of great significance for the early diagnosis of BC. The level of its serum content can be used as an index for the evaluation of the efficacy of BC^[24, 25]^, it is currently clinically recognized as a more specific tumor marker for the diagnosis of BC. In this study, we found that the serum CA153 level was significantly higher in Luminal subtype patients than in non-Luminal subtype patients. There fore, we hypothesized that the serum CA153 difference in different BC subtype s combined with CEUS related parameters could help us to better predict the molecular subtype of BC. Based on this, we used multivariate Logistic regression to analyze the clinical data, contrast-enhanced ultrasound characteristic para meters and serum CA153 of the two groups of patients. The results showed th at the quantitative parameters of contrast-enhanced ultrasound PI, AUC and serum CA153 were independent predictors of BC molecular typing. The ROC curve was used to compare the efficacy of PI, AUC, CA153 single parameters an d the combination of three parameters in the diagnosis of BC molecular typing, and the results showed that the AUC of the combination of three parameters was significantly higher than that of each parameter. Then we constructed a nomogram prediction model using three parameters, and evaluated the predictive performance of the model using ROC curve and calibration curve. The results of ROC curve and calibration curve showed that the nomogram had good predictive value for the risk of non-Luminal BC.

In conclusion, the ultrasonographic features of BC can reflect the biological characteristics of different BC molecular typing and are closely associated wit h the serum tumor marker CA153. In this study, the ultrasonographic parameters of PI, AUC and serum CA153 were used to construct a columnar graphical model to effectively predict the molecular typing of BC, which can provide certain clinical references for accurate diagnosis and personalized treatment of BC. However, this study is a single-center retrospective analysis with a limited sam ple size, which did not explore the specific molecular typing of BC and lacked external data validation, so it needs to be further improved.

## Data Availability

NO

## Author contributions

Yanlei Ji, Zhen Han, and Yong Zhang: study design, data analysis and interpretation, and manuscript revision. Wenwen Sun: manuscript drafing. All the authors of this study read and approved the fnal manuscript.

